# Disease severity of unvaccinated SARS-CoV-2 positive adults less than 65 years old without comorbidity, in the Omicron period and pre-Omicron periods

**DOI:** 10.1101/2023.02.02.23285377

**Authors:** Erik Wahlström, Daniel Bruce, Anna M Bennet-Bark, Sten Walther, Håkan Hanberger, Kristoffer Strålin

## Abstract

**Background:** The reduced severity and burden of COVID-19 in 2022 can largely be attributable to vaccination and a shift to Omicron predominance. However, millions of individuals remain unvaccinated. In the present study, we aimed to study disease severity in unvaccinated individuals without risk factors during the Omicron period, compared to pre-Omicron periods.

**Methods:** This register-based study included all unvaccinated individuals in Sweden aged 18-64 years without comorbidity or care dependency who were SARS-CoV-2 positive between week 45 of 2020 and week 5 of 2022. Variant of concern (VOC) periods were periods with certain VOCs identified in ≥92% of sequenced cases nationwide. Outcomes were hospitalization with a main discharge code of COVID-19; severe illness, defined as high-flow nasal oxygen treatment or intensive care unit admission; and death with COVID-19 as the underlying cause of death on the death certificate.

**Results:** Among 788,895 individuals in the overall SARS-CoV-2 positive cohort, both hospitalization and death increased stepwise from the pre-VOC period to the Alpha and Delta periods, and decreased in the Omicron period. Among 15,179 patients hospitalized for COVID-19, the proportions with severe illness and death increased to the Delta period, but in the Omicron period, these outcomes returned to the level of the pre-VOC period.

**Conclusion:** In the Omicron period, compared to pre-Omicron periods, unvaccinated SARS-CoV-2 positive adults <65 years old without comorbidity had reduced proportions of hospitalization and death overall, but similar proportion of severe illness among patients hospitalized for COVID-19. These results support continuous efforts to prevent hospitalizations for COVID-19.

## Introduction

The healthcare burden of COVID-19 in Western countries decreased in 2022, most likely due to the combination of widespread vaccination [1] and a shift to the Omicron SARS-CoV-2 variant of concern (VOC) [2]. However, millions of individuals remain unvaccinated [3], and the severity of COVID-19 in this population during the Omicron period has not been fully clarified. Thus, in a nationwide register-based study, we aimed to study the disease severity among unvaccinated SARS-CoV-2-positive individuals over time, comparing the Omicron period to pre-Omicron periods. As advanced age, comorbidity, and care dependency have a great impact on disease severity and outcomes from COVID-19, the present study was focused on adult individuals < 65 years of age without comorbidity or care dependency.

## Methods

### Population

The study included all unvaccinated individuals in Sweden aged 18-64 years without comorbidity or care dependency with a first-time SARS-CoV-2-positive test from samples collected between week 45 of 2020 and week 5 of 2022, in Sweden. The national recommendations for public testing were unaltered during this period up to week 3 of 2022. In weeks 4 and 5 of 2022, some limitations to the public testing were introduced, and from week 6 the public testing was withdrawn [4].

Nationwide registry data were compiled by the Swedish National Board of Health and Welfare using the unique national personal identification number as described previously [5, 6]. Briefly, data on demographic factors, comorbidity diagnoses, drugs dispensed during the last 5 years (for comorbidity information), care dependency, hospitalizations, discharge codes, mortality, and death certificates were obtained from the registers of the National Board of Health and Welfare. Anti-SARS-CoV-2 vaccination data were obtained from the Public Health Agency of Sweden. Intensive care unit (ICU) data were obtained from the Swedish Intensive Care Registry. Socio-economic data (country of birth, education level, disposable income, main source of income) were obtained from the Swedish Longitudinal Integrated Database for Health Insurance and Labour Market Studies held by Statistics Sweden.

### Outcomes

Outcome measures were hospitalization for COVID-19, defined as hospitalization within 14 days after to 5 days before the first SARS-CoV-2-positive test combined with a main discharge code of COVID-19; severe illness, defined as either treatment with high-flow nasal oxygen (according to a specific national discharge code) or admission to an ICU during hospitalization for COVID-19; invasive mechanical ventilation during hospitalization for COVID-19; and death from COVID-19, defined as death within 60 days from SARS-CoV-2 positivity with COVID-19 specified as the underlying cause of death on the death certificate.

### Variant of concern periods

Aggregated data on the distributions of each VOC per week based on all sequenced SARS-CoV-2-viruses nationally were available as open data from the Public Health Agency of Sweden [7]. VOC periods were defined as weeks in which a certain SARS-CoV-2 VOC constituted ≥92% of all whole genome-sequenced SARS-CoV-2-positive cases in Sweden. Weeks with <92% of a certain VOC were considered mix periods. This threshold was selected based on national whole genome sequence data to enable an adequately large Alpha cohort while achieving an Omicron period with limited influence from cases infected with Delta VOC. In the first weeks of 2021, the proportions of VOCs among SARS-CoV-2-positive cases were uncertain due to limited sequencing of samples. The Pre-VOC period was defined as the period prior to week 52 of 2020, when only sporadic cases of Alpha VOC had been identified [7].

### Statistical analysis

Outcomes are presented as absolute proportions (with 95% confidence intervals; CIs) and adjusted odds ratios (ORs) with 95% CIs using the Pre-VOC period as a reference.

ORs for study outcomes (hospitalization and death after SARS-CoV-2-positivity and severe illness, invasive mechanical ventilation, and death after hospitalization) were estimated using logistic regression models with VOC period as exposure variable of interest. The estimates were adjusted for age (continuous; both a linear and quadratic term), sex (categorical; male/female), country of birth (categorical; Africa/Asia/Central and Eastern Europe/Middle East/Scandinavia/Sweden/South America/Southeast Europe/Other), income quintile (categorical; by quintiles of the distribution in the total population), income source (categorical, categorized as ‘employed/student/caregiver’, ‘disability pension’, ‘sick pay/unemployment’, ‘retirement pension’, ‘financial aid’), and education (categorical; primary school, secondary school, university/college < 3 years, university/college > 3 years), all modelled as main effects.

In all regression models, missing data were handled by complete case analysis. Data were complete on all variables except for country of birth (2% missing), education (3%), income (4%), and income source (5%).

All data management and statistical analyses was performed using SAS Software SAS Enterprise Guide version 8.3 (SAS institute, Cary, NC, USA).

### Ethical permission

The study was approved by the Swedish Ethics Review Authority, Uppsala (Dnr 2020-04278).

## Results

### Variant of concern periods

Supplementary Fig. S1A shows the distribution of SARS-CoV-2 VOCs over time among whole genome-sequenced cases in Sweden. Using the threshold of 92% among these whole genome-sequenced cases to define a VOC period, the following VOC periods and mix periods were identified for the present study: Alpha period, weeks 14 – 21 of 2021; Mix Alpha-Delta period, weeks 22 – 27 of 2021; Delta period, weeks 28 – 49 of 2021; Mix Delta-Omicron period, week 50 of 2021 – week 1 of 2022; and Omicron period, weeks 2 – 5 of 2022. In addition, we included a Pre-VOC period, weeks 45 – 51 of 2020, when only sporadic cases of VOCs were identified, and a Mix Pre-VOC-Alpha period, week 52 of 2020 – week 13 of 2021.

### Population and study cases

Among all SARS-CoV-2-positive individuals in Sweden aged 18-64 years without comorbidity or care dependency, the proportion of unvaccinated and first-time positive individuals (i.e., the overall cohort of the present study) decreased from 100% in the Pre-VOC period to <20% during the Omicron period (Suppl. Fig. S2).

Numbers of study cases per week are shown in Supplementary Fig. S1B (overall cohort) and Supplementary Fig. S1C (hospitalized cohort, i.e., cases hospitalized for COVID-19).

### Characteristics and outcomes in the overall cohort

A total of 788,895 unvaccinated first-time SARS-CoV-2-positive individuals aged 18-64 years without comorbidity or care dependency were included in this analysis. The characteristics of the study population changed over time (Table 1). The populations of the Pre-VOC period, Alpha period, Delta period, and Omicron period were 53%, 48%, 49%, and 51% female with a median age of 40 years, 38 years, 32 years, and 35 years, respectively. The proportions of these populations born outside of Sweden were 27%, 22%, 47%, and 42%, respectively, and 13%, 13%, 26%, and 24% belonged to the lowest income quintile.

**Table 1.** Characteristics of study individuals in the overall cohort.

| Characteristics | All individuals<br>(n = 788895)<br>No. (%) | SARS-CoV-2 variant predominance period |  |  |  |  |  |  |
| --- | --- | --- | --- | --- | --- | --- | --- | --- |
|  |  | Pre-VOC<br>(n = 179856)<br>No. (%) | Mix Pre-VOC<br>– Alpha<br>(n = 310866)<br>No. (%) | Alpha<br>(n = 157070)<br>No. (%) | Mix Alpha<br>– Delta<br>(n = 11346)<br>No. (%) | Delta<br>(n = 35188)<br>No. (%) | Mix Delta –<br>Omicron<br>(n = 27415)<br>No. (%) | Omicron<br>(n = 67154)<br>No. (%) |
| **Sex** |  |  |  |  |  |  |  |  |
| Male | 392886 (50) | 84815 (47) | 155979 (50) | 81880 (52) | 5968 (53) | 17860 (51) | 13679 (50) | 32705 (49) |
| Female | 396009 (50) | 95041 (53) | 154887 (50) | 75190 (48) | 5378 (47) | 17328 (49) | 13736 (50) | 34449 (51) |
| **Age** |  |  |  |  |  |  |  |  |
| 18-34 years | 327521 (42) | 68875 (38) | 120368 (39) | 64898 (41) | 6872 (61) | 20254 (58) | 14542 (53) | 31712 (47) |
| 35-49 years | 276199 (35) | 62694 (35) | 108114 (35) | 55682 (35) | 3522 (31) | 11222 (32) | 9265 (34) | 25700 (38) |
| 50-64 years | 185175 (23) | 48287 (27) | 82384 (27) | 36490 (23) | 952 (8.4) | 3712 (11) | 3608 (13) | 9742 (15) |
| **Region of birth** |  |  |  |  |  |  |  |  |
| Africa | 22238 (2.8) | 4759 (2.6) | 6817 (2.2) | 3161 (2.0) | 345 (3.0) | 2225 (6.3) | 1635 (6.0) | 3296 (4.9) |
| Asia | 26606 (3.4) | 6364 (3.5) | 10762 (3.5) | 5261 (3.3) | 444 (3.9) | 1201 (3.4) | 765 (2.8) | 1809 (2.7) |
| Central and east Europe | 20583 (2.6) | 3266 (1.8) | 5861 (1.9) | 2891 (1.8) | 195 (1.7) | 2091 (5.9) | 1495 (5.5) | 4784 (7.1) |
| Middle East | 65502 (8.3) | 16060 (8.9) | 24187 (7.8) | 10477 (6.7) | 858 (7.6) | 4801 (14) | 2507 (9.1) | 6612 (9.8) |
| Nordic countries (not Sweden) | 10092 (1.3) | 2415 (1.3) | 4004 (1.3) | 1900 (1.2) | 103 (0.91) | 381 (1.1) | 377 (1.4) | 912 (1.4) |
| Sweden | 565377 (72) | 131295 (73) | 232747 (75) | 120929 (77) | 8632 (76) | 18221 (52) | 16007 (58) | 37546 (56) |
| South America | 9733 (1.2) | 2436 (1.4) | 3578 (1.2) | 1793 (1.1) | 115 (1.0) | 476 (1.4) | 401 (1.5) | 934 (1.4) |
| Southeast Europe | 42206 (5.4) | 8428 (4.7) | 14616 (4.7) | 6078 (3.9) | 268 (2.4) | 3848 (11) | 2355 (8.6) | 6613 (9.8) |
| Other | 11854 (1.5) | 2778 (1.5) | 4361 (1.4) | 2267 (1.4) | 148 (1.3) | 519 (1.5) | 491 (1.8) | 1290 (1.9) |
| Missing data | 14704 (1.9) | 2055 (1.1) | 3933 (1.3) | 2313 (1.5) | 238 (2.1) | 1425 (4.0) | 1382 (5.0) | 3358 (5.0) |
| **Income quintile** |  |  |  |  |  |  |  |  |
| 1st | 117922 (15) | 23463 (13) | 40654 (13) | 20947 (13) | 1848 (16) | 9007 (26) | 6114 (22) | 15889 (24) |
| 2nd | 136003 (17) | 28710 (16) | 51023 (16) | 25902 (16) | 2033 (18) | 7559 (21) | 5636 (21) | 15140 (23) |
| 3rd | 166192 (21) | 37209 (21) | 66474 (21) | 33893 (22) | 2313 (20) | 6954 (20) | 5450 (20) | 13899 (21) |
| 4th | 178949 (23) | 42841 (24) | 74248 (24) | 38006 (24) | 2580 (23) | 5592 (16) | 4812 (18) | 10870 (16) |
| 5th | 163402 (21) | 43682 (24) | 70840 (23) | 33903 (22) | 2138 (19) | 3611 (10) | 2996 (11) | 6232 (9.3) |
| Missing data | 26427 (3.3) | 3951 (2.2) | 7627 (2.5) | 4419 (2.8) | 434 (3.8) | 2465 (7.0) | 2407 (8.8) | 5124 (7.6) |
| **Main income source** |  |  |  |  |  |  |  |  |
| Disability pension | 9774 (1.2) | 2215 (1.2) | 4150 (1.3) | 1685 (1.1) | 77 (0.68) | 388 (1.1) | 340 (1.2) | 919 (1.4) |
| Employed, student, or parental | 705399 (89) | 165012 (92) | 282863 (91) | 142580 (91) | 10204 (90) | 28973 (82) | 21957 (80) | 53810 (80) |
| Financial aid | 17082 (2.2) | 3319 (1.8) | 5745 (1.8) | 3030 (1.9) | 287 (2.5) | 1512 (4.3) | 845 (3.1) | 2344 (3.5) |
| Retired | 1935 (0.25) | 565 (0.31) | 823 (0.26) | 346 (0.22) | 26 (0.23) | 63 (0.18) | 35 (0.13) | 77 (0.11) |
| Sickpay or unemployed | 21025 (2.7) | 4259 (2.4) | 7583 (2.4) | 3811 (2.4) | 242 (2.1) | 1401 (4.0) | 1045 (3.8) | 2684 (4.0) |
| Missing data | 33680 (4.3) | 4486 (2.5) | 9702 (3.1) | 5618 (3.6) | 510 (4.5) | 2851 (8.1) | 3193 (12) | 7320 (11) |
| **Education** |  |  |  |  |  |  |  |  |
| Primary school | 111501 (14) | 19401 (11) | 40160 (13) | 24001 (15) | 2311 (20) | 7685 (22) | 4925 (18) | 13018 (19) |
| Secondary school | 354029 (45) | 78863 (44) | 141323 (45) | 69879 (44) | 4801 (42) | 15652 (44) | 12789 (47) | 30722 (46) |
| Tertiary school less than 3 | 116894 (15) | 27917 (16) | 47572 (15) | 23221 (15) | 1635 (14) | 4623 (13) | 3597 (13) | 8329 (12) |
| Tertiary school 3 or more | 185663 (24) | 50615 (28) | 76002 (24) | 36936 (24) | 2280 (20) | 5209 (15) | 4060 (15) | 10561 (16) |
| Missing data | 20808 (2.6) | 3060 (1.7) | 5809 (1.9) | 3033 (1.9) | 319 (2.8) | 2019 (5.7) | 2044 (7.5) | 4524 (6.7) |

Abbreviations: VOC, variant of concern.

Fig. 1A shows gradual increases in the proportion of hospitalizations for COVID-19, from 1.6% in the Pre-VOC period to 3.9% in the Delta period, followed by a decrease to 0.3% in the Omicron period. Similar patterns were noted in all age subgroups (18-34 years, 35-49 years, 50-64 years), though the proportion hospitalized was significantly higher for those aged 50-64 years compared to those aged 18-49 years, in all periods (Fig. 2, left panel). With the Pre-VOC period as a reference, the adjusted ORs for hospitalization for COVID-19 were 1.74 (95% CI, 1.65-1.83) for the Alpha period, 3.14 (95% CI, 2.91-3.38) for the Delta period, and 0.30 (95% CI, 0.26-0.34) for the Omicron period (Fig. 1B).

**Fig. 1.**
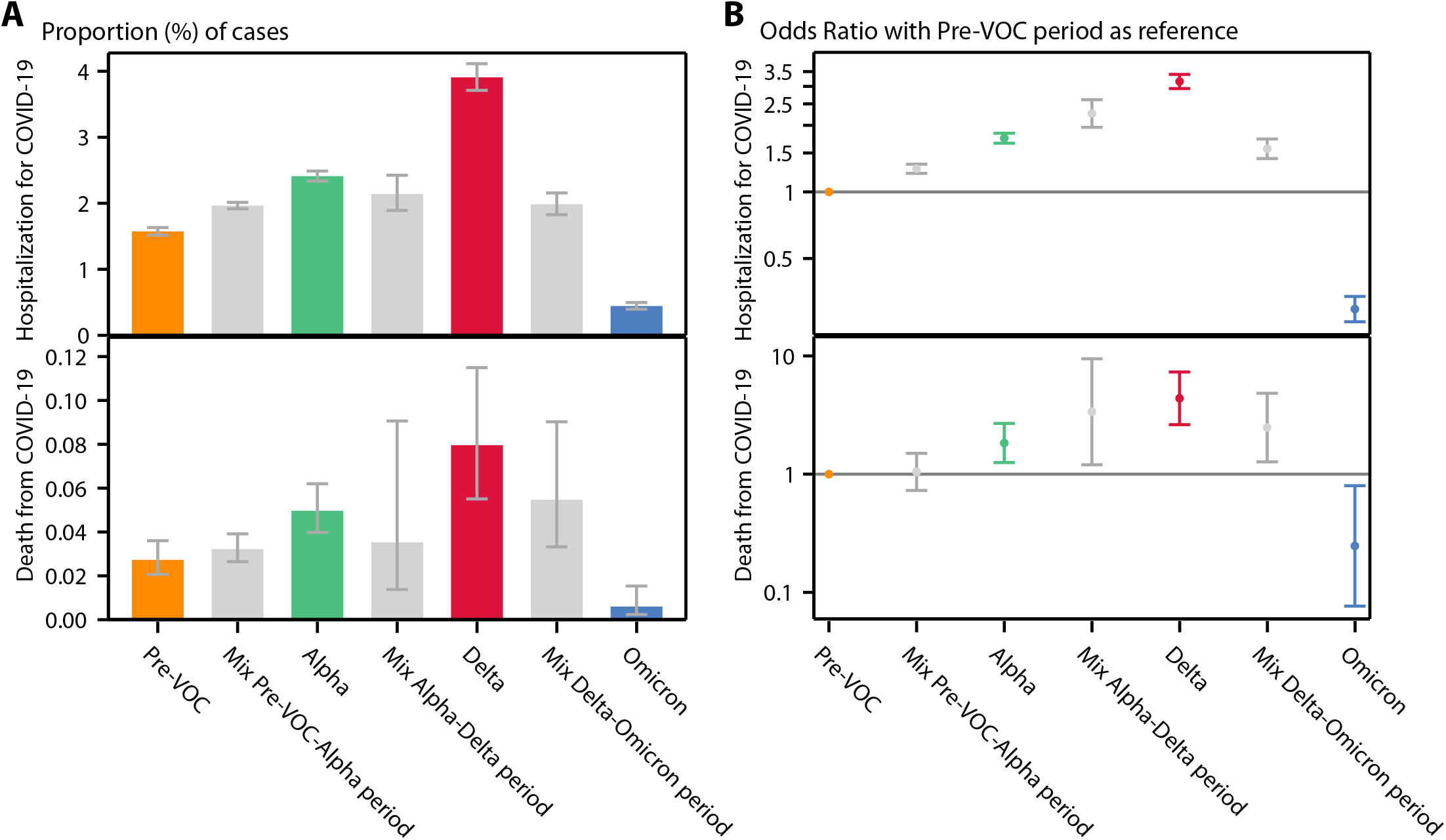
A) Proportions of study outcomes with 95% confidence intervals according to VOC predominance in the overall cohort. B) Odds ratios for study outcomes with 95% confidence intervals adjusted for age, sex, and socio-economic status with the Pre-VOC period as reference in the overall cohort.

**Fig. 2.**
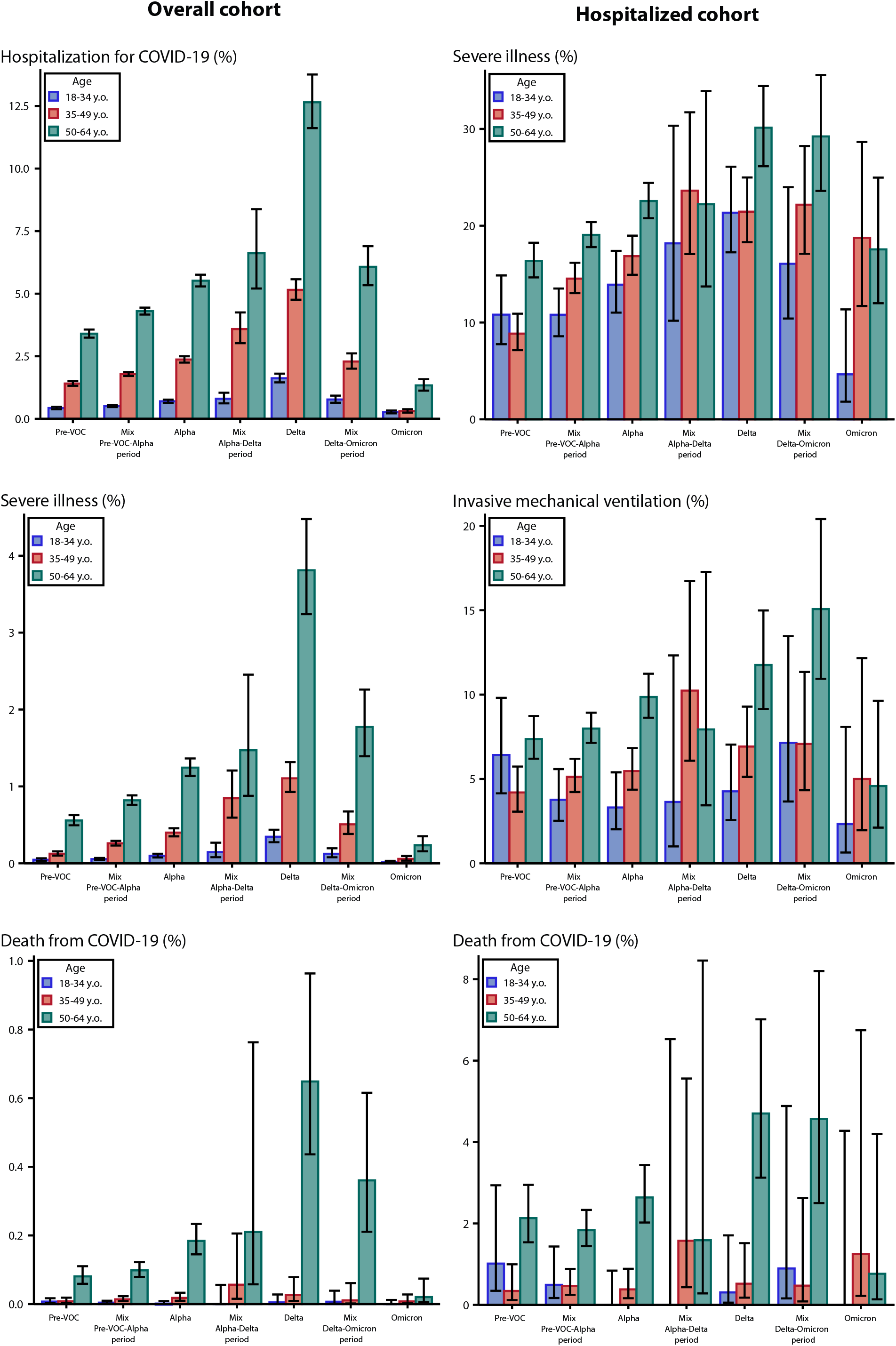
Proportions of study outcomes with 95% confidence intervals according to age group in different SARS-CoV-2 variant predominance periods. Left panel, outcomes of the overall cohort; Right panel, outcomes of the hospitalized cohort, i.e. patients hospitalized for COVID-19.

Similarly, death from COVID-19 increased from 0.027% in the Pre-VOC period to 0.079% in the Delta period and decreased to 0.006% in the Omicron period. As noted in Fig. 2 (left panel), the proportion with death from COVID-19 in the overall cohort was dramatically higher in individuals aged 50-64 years than in those aged 18-49, throughout the study period, and a similar pattern was noted for severe illness. Overall, the adjusted ORs for death from COVID-19 were 1.84 (95% CI, 1.26-2.69) for the Alpha period, 4.39 (95% CI, 2.63-7.33) for the Delta period, and 0.25 (95% CI, 0.077-0.81) for the Omicron period (Fig. 1B).

### Characteristics and outcomes in the hospitalized cohort

A total of 15,179 unvaccinated first-time SARS-CoV-2-positive adults aged 18-64 years without comorbidity or care dependency were hospitalized for COVID-19 with a main discharge code of COVID-19 and included in this analysis. The characteristics of the study population changed over time (Table 2). The populations of the Pre-VOC period, Alpha period, Delta period, and Omicron period were 38%, 38%, 40%, and 50% female with a median age of 52 years, 50 years, 44 years, and 46 years, respectively. The proportions of these populations born outside of Sweden were 50%, 41%, 67%, and 59%, respectively, and 22%, 19%, 34%, and 40% belonged to the lowest income quintile.

**Table 2.** Characteristics of study patients hospitalized for COVID-19.

| Characteristics | All patients<br>(n = 15179)<br>No. (%) | SARS-CoV-2 variant predominance period |  |  |  |  |  |  |
| --- | --- | --- | --- | --- | --- | --- | --- | --- |
|  |  | Pre-VOC<br>(n = 2829)<br>No. (%) | Mix Pre-VOC<br>– Alpha<br>(n = 6105)<br>No. (%) | Alpha<br>(n = 3786)<br>No. (%) | Mix Alpha<br>– Delta<br>(n = 243)<br>No. (%) | Delta<br>(n = 1375)<br>No. (%) | Mix Delta –<br>Omicron<br>(n = 544)<br>No. (%) | Omicron<br>(n = 297)<br>No. (%) |
| Sex |  |  |  |  |  |  |  |  |
| Male | 9417 (62) | 1767 (62) | 3852 (63) | 2355 (62) | 150 (62) | 824 (60) | 321 (59) | 148 (50) |
| Female | 5762 (38) | 1062 (38) | 2253 (37) | 1431 (38) | 93 (38) | 551 (40) | 223 (41) | 149 (50) |
| Age |  |  |  |  |  |  |  |  |
| 18-34 years | 1952 (13) | 302 (11) | 615 (10) | 454 (12) | 54 (22) | 329 (24) | 112 (21) | 86 (29) |
| 35-49 years | 5136 (34) | 883 (31) | 1938 (32) | 1320 (35) | 125 (51) | 577 (42) | 213 (39) | 80 (27) |
| 50-64 years | 8091 (53) | 1644 (58) | 3552 (58) | 2012 (53) | 64 (26) | 469 (34) | 219 (40) | 131 (44) |
| Region of birth |  |  |  |  |  |  |  |  |
| Africa | 646 (4.3) | 129 (4.6) | 222 (3.6) | 139 (3.7) | 7 (2.9) | 94 (6.8) | 33 (6.1) | 22 (7.4) |
| Asia | 989 (6.5) | 193 (6.8) | 408 (6.7) | 253 (6.7) | 27 (11) | 72 (5.2) | 28 (5.1) | 8 (2.7) |
| Central and east Europe | 400 (2.6) | 48 (1.7) | 122 (2.0) | 68 (1.8) | 8 (3.3) | 99 (7.2) | 39 (7.2) | 16 (5.4) |
| Middle East | 2688 (18) | 553 (20) | 1004 (16) | 569 (15) | 58 (24) | 344 (25) | 88 (16) | 72 (24) |
| Nordic countries (not Sweden) | 337 (2.2) | 60 (2.1) | 139 (2.3) | 91 (2.4) | 5 (2.1) | 24 (1.7) | 12 (2.2) | 6 (2.0) |
| Sweden | 7865 (52) | 1396 (49) | 3368 (55) | 2205 (58) | 114 (47) | 429 (31) | 233 (43) | 120 (40) |
| South America | 363 (2.4) | 87 (3.1) | 124 (2.0) | 93 (2.5) | 3 (1.2) | 33 (2.4) | 19 (3.5) | 4 (1.3) |
| Southeast Europe | 1432 (9.4) | 276 (9.8) | 563 (9.2) | 276 (7.3) | 10 (4.1) | 207 (15) | 62 (11) | 38 (13) |
| Other | 178 (1.2) | 33 (1.2) | 73 (1.2) | 45 (1.2) | 2 (0.82) | 17 (1.2) | 7 (1.3) | 1 (0.34) |
| Missing data | 281 (1.9) | 54 (1.9) | 82 (1.3) | 47 (1.2) | 9 (3.7) | 56 (4.1) | 23 (4.2) | 10 (3.4) |
| Income quintile |  |  |  |  |  |  |  |  |
| 1st | 3345 (22) | 623 (22) | 1203 (20) | 703 (19) | 62 (26) | 464 (34) | 172 (32) | 118 (40) |
| 2nd | 2513 (17) | 436 (15) | 981 (16) | 599 (16) | 50 (21) | 275 (20) | 117 (22) | 55 (19) |
| 3rd | 2864 (19) | 498 (18) | 1170 (19) | 748 (20) | 44 (18) | 243 (18) | 109 (20) | 52 (18) |
| 4th | 2976 (20) | 550 (19) | 1263 (21) | 831 (22) | 43 (18) | 186 (14) | 67 (12) | 36 (12) |
| 5th | 2919 (19) | 633 (22) | 1307 (21) | 790 (21) | 25 (10) | 107 (7.8) | 41 (7.5) | 16 (5.4) |
| Missing data | 562 (3.7) | 89 (3.1) | 181 (3.0) | 115 (3.0) | 19 (7.8) | 100 (7.3) | 38 (7.0) | 20 (6.7) |
| Main income source |  |  |  |  |  |  |  |  |
| Disability pension | 880 (5.8) | 190 (6.7) | 384 (6.3) | 168 (4.4) | 5 (2.1) | 66 (4.8) | 31 (5.7) | 36 (12) |
| Employed, student, or parental | 11713 (77) | 2181 (77) | 4769 (78) | 3061 (81) | 190 (78) | 955 (69) | 379 (70) | 178 (60) |
| Financial aid | 810 (5.3) | 156 (5.5) | 280 (4.6) | 178 (4.7) | 12 (4.9) | 112 (8.1) | 47 (8.6) | 25 (8.4) |
| Retired | 95 (0.63) | 26 (0.92) | 43 (0.70) | 19 (0.50) | 0 | 5 (0.36) | 1 (0.18) | 1 (0.34) |
| Sickpay or unemployed | 934 (6.2) | 158 (5.6) | 374 (6.1) | 211 (5.6) | 17 (7.0) | 113 (8.2) | 36 (6.6) | 25 (8.4) |
| Missing data | 747 (4.9) | 118 (4.2) | 255 (4.2) | 149 (3.9) | 19 (7.8) | 124 (9.0) | 50 (9.2) | 32 (11) |
| Education |  |  |  |  |  |  |  |  |
| Primary school | 2769 (18) | 523 (18) | 1067 (17) | 618 (16) | 41 (17) | 311 (23) | 127 (23) | 82 (28) |
| Secondary school | 6743 (44) | 1177 (42) | 2755 (45) | 1736 (46) | 97 (40) | 603 (44) | 253 (47) | 122 (41) |
| Tertiary school less than 3 | 2151 (14) | 408 (14) | 893 (15) | 562 (15) | 38 (16) | 157 (11) | 59 (11) | 34 (11) |
| Tertiary school 3 or more | 3011 (20) | 617 (22) | 1230 (20) | 774 (20) | 52 (21) | 232 (17) | 71 (13) | 35 (12) |
| Missing data | 505 (3.3) | 104 (3.7) | 160 (2.6) | 96 (2.5) | 15 (6.2) | 72 (5.2) | 34 (6.3) | 24 (8.1) |

Abbreviations: VOC, variant of concern.

Fig. 3A shows a gradual increase in severe illness, from 13% in the Pre-VOC period to 23% in the Delta period, followed by a decrease to 14% in the Omicron period. The patients with severe illness in the Omicron period had a median age of 53 years (interquartile range, 39-63 years), 38% were female. Similar patterns over time were noted in all age subgroups (18-34 years, 35-49 years, 50-64 years; Fig. 2, right panel). However, the proportion of severe illness in the Omicron period was lower in patients aged 18-34 years (4%) than in patients aged 35-49 years (19%) and 50-64 years (18%). In the adjusted analysis with the Pre-VOC period as a reference (Fig. 3B), there was a stepwise increase in severe illness to the Alpha period (OR 1.53; 95% CI, 1.33-1.76) and Delta period (OR 2.37; 95% CI, 1.98-2.84), followed by a decrease in the Omicron period (OR 1.22; 95% CI, 0.84-1.76).

**Fig. 3.**
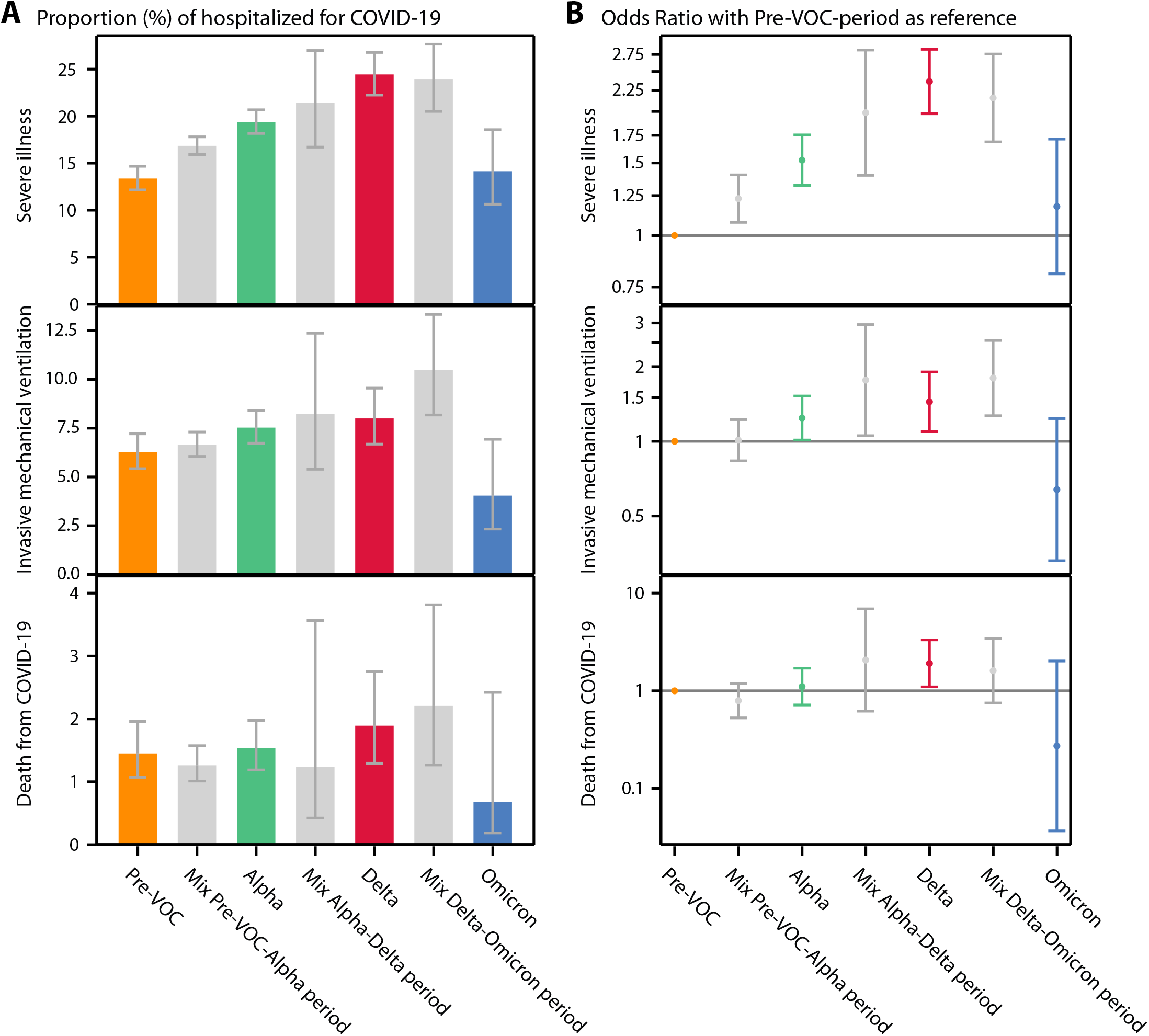
A) Proportions of study outcomes among patients hospitalized for COVID-19 with 95% confidence intervals according to VOC predominance. B) Odds ratios for study outcomes among patients hospitalized for COVID-19 with 95% confidence intervals adjusted for age, sex, and socio-economic status with the Pre-VOC period as reference.

In the hospitalized cohort, the proportion of patients with invasive mechanical ventilation and death from COVID-19 did not significantly change over time (Fig. 3A). However, in the adjusted analysis with the Pre-VOC period as a reference (Fig. 3B), higher ORs for invasive mechanical ventilation were noted for both the Alpha period (OR 1.24; 95% CI, 1.01-1.52) and Delta period (OR 1.44; 95% CI, 1.10-1.90), and a higher OR for death from COVID-19 was noted for the Delta period (OR 1.92; 95% CI, 1.10-3.34). With the Delta period used as a reference in a complementary analysis of patients hospitalized for COVID-19, the Omicron period had adjusted ORs of 0.5 (95% CI, 0.3-0.7) for severe illness, 0.45 (95% CI, 0.20-0.75) for invasive mechanical ventilation, and 0.1 (95% CI, 0.02-1.05) for death from COVID-19 (Suppl. Fig. S3).

## Discussion

The overall study clearly shows that, among unvaccinated first-time SARS-CoV-2-positive adults aged < 65 years without comorbidity or care dependency, the proportion with hospitalization for COVID-19 and death from COVID-19 increased from the Pre-VOC period to the Delta period, but then dropped considerably with the Omicron period. Among patients hospitalized for COVID-19, the proportion with severe illness and death from COVID-19 increased from the Pre-VOC to the Delta period but, in the Omicron period, these outcomes returned to the level of the Pre-VOC period.

The study results are in agreement with previous studies indicating that the Alpha VOC causes more severe disease than non-VOC virus [5, 8], that the Delta VOC causes more severe disease than the Alpha VOC [9], and that the Omicron VOC causes milder disease than the Delta VOC [2, 10-14]. Importantly, the present study adds an understanding of the magnitude of changes in severity between different periods of VOC predominance, as the population was not affected by old age, comorbidity, previous infection, or vaccination.

Compared to the Pre-VOC period in the overall cohort, there was a stepwise increase in severity to the Delta period, with ORs of > 3 for hospitalization and > 4 for death from COVID-19, followed by a drop in severity in the Omicron period, with ORs of 0.3 for hospitalization and 0.25 for death from COVID-19 (Fig. 1B). Among hospitalized patients, compared to the Pre-VOC period, severity increased to the Delta period, with ORs of approximately 2 for both severe illness and death from COVID-19, but in the Omicron period, severity returned to the level of the Pre-VOC period (Fig. 3B). Compared to the Delta period, hospitalized patients in the Omicron period showed decreased severity, with ORs of approximately 0.5 for both severe illness and invasive mechanical ventilation (Suppl. Fig. S3). The severe illness among hospitalized patients in the Omicron period was mainly driven by patients aged 35-64 years, as those aged 18-34 had low proportion of severe illness (Fig. 2, right panel).

The study findings are in line with those of Lauring et al. [15], who found that unvaccinated hospitalized COVID-19 patients had substantial disease severity in the Omicron period; the disease severity was reported to be 79% compared to that of Alpha and 61% compared to that of Delta. Accordingly, Robinson et al. [16] found that the relative risk of severe disease or death among unvaccinated hospitalized COVID-19 patients was similar between cases with Omicron and cases with ancestral lineages of SARS-CoV-2 virus. This documented disease severity of unvaccinated patients requiring hospitalization for COVID-19 in the Omicron period motivates continuous efforts to prevent hospitalizations for SARS-CoV-2 infection, mainly through vaccination and optimized use of early outpatient antiviral treatment.

The present study illustrates how the clinical course of COVID-19 has changed since the shift to Omicron VOC predominance. The reduced proportion of infected cases requiring hospitalization in the Omicron period is encouraging. However, the enhanced transmissibility of the Omicron virus is concerning, as loads of individuals may be infected at the same time, thus causing considerable numbers of patients requiring hospital care. There is a need for continuous surveillance of the evolution of the SARS-CoV-2 virus, both regarding its transmissibility and its link to disease severity.

This study has a number of strengths. First, the exclusion of individuals with old age and comorbidities enabled a pure analysis of the changes over time in unvaccinated first-time positive individuals. Second, the large sample size enabled analysis of both an overall national cohort and a cohort of patients hospitalized for COVID-19. Third, during the study period, no major changes occurred in the public sampling strategy towards COVID-19. Fourth, during the study period, no major changes occurred in recommendations regarding medical treatment and respiratory treatment for COVID-19 in Sweden.

This study also has a number of limitations. First, the patients’ individual SARS-CoV-2 variants were not determined. However, we used nationwide data on whole genome-sequenced SARS-CoV-2 cases and required at least 92% dominance to define a VOC period. Second, the gradually reduced number of unvaccinated individuals and changing characteristics of the study population may have influenced the results. However, the changes remained after adjusting for age, sex, and socio-economic status. Third, the strain on hospitals varied over time and may have led to varying thresholds to admit individuals with mild COVID-19. However, the threshold to treat patients without risk factors with high-flow nasal oxygen or to admit them to an ICU most likely was not influenced by COVID-19 hospital strain. Forth, the study results may have been influenced by undiagnosed SARS-CoV-2 infection in the first pandemic wave when there was no public testing. However, the seroprevalence among unvaccinated individuals in Sweden was reported to be low (20%) after the first year of the pandemic [17].

In conclusion, among unvaccinated first-time SARS-CoV-2-positive individuals without risk factors, disease severity gradually increased from the Pre-VOC period to the Delta period, and then decreased with the Omicron period. However, patients hospitalized for COVID-19 during the Omicron period had disease severity similar to that of the Pre-VOC period. The results motivate continuous efforts to prevent hospitalization for COVID-19, mainly through vaccination, and continuous surveillance of virus evolution and its link to disease severity.

## Data Availability

The data underlying this article cannot be shared publicly due to regulations under Swedish law. According to the Swedish Ethics Review Act, the General Data Protection Regulation, and the Public Access to Information and Secrecy Act, patient data can only be made available, after legal review, to researchers who meet the criteria for access to this type of confidential data. Requests regarding data in this report may be made to the corresponding author.

## Acknowledgments

The authors thank Johanna Holm for critical review of the manuscript.

## References

1. Haas EJ, Angulo FJ, McLaughlin JM, et al. Impact and effectiveness of mRNA BNT162b2 vaccine against SARS-CoV-2 infections and COVID-19 cases, hospitalisations, and deaths following a nationwide vaccination campaign in Israel: an observational study using national surveillance data. Lancet 2021; 397:1819–29.

2. Lewnard JA, Hong VX, Patel MM, et al. Clinical outcomes associated with SARS-CoV-2 Omicron (B.1.1.529) variant and BA.1/BA.1.1 or BA.2 subvariant infection in southern California. Nat Med 2022; 28:1933–43.

3. Zamir E, Gillis P. The pandemic of the unvaccinated: a Covid-19 ethical dilemma. Heart Lung 2023; 57:292–4..

4. Public Health Agency of Sweden. Veckorapport om covid-19, vecka 6. 2022. Available at: https://www.folkhalsomyndigheten.se/globalassets/statistik-uppfoljning/smittsamma-sjukdomar/veckorapporter-covid-19/2022/covid-19-veckorapport-2022-vecka-6_uppdaterad-20-april-2022.pdf. Accessed 12 January 2023.

5. Strålin K, Bruce D,Wahlström E,,et al. Impact of the Alpha VOC on disease severity in SARS-CoV-2-positive adults in Sweden. J Infect 2022; 84:e3–e5..

6. Strålin K, Wahlström E, Walther S, et al. Mortality in hospitalized COVID-19 patients was associated with the COVID-19 admission rate during the first year of the pandemic in Sweden. Infect Dis (Lond.) 2022; 54:145–41.

7. Public Health Agency of Sweden. Helgenomsekvensering av svenska SARS-CoV-2 som orsakar covid-19, del 4. 2022. Available at: https://www.folkhalsomyndigheten.se/contentassets/d0dcd4715e284db4a9ecbf52d9784e0d/helgenomsekvensering-svenska-sars-cov-2-rapport-4.pdf. Accessed 12 January 2023.

8. Cevik M, Mishra S. SARS-CoV-2 variants and considerations of inferring causality on disease severity. Lancet Infect Dis 2021; 21:1472–4.

9. Twohig KA, Nyberg T, Zaidi A, et al. Hospital admission and emergency care attendance risk for SARS-CoV-2 delta (B.1.617.2) compared with alpha (B.1.1.7) variants of concern: a cohort study. Lancet Infect Dis 2022; 22:35–42.

10. Bager P, Wohlfahrt J, Bhatt S, et al. Risk of hospitalisation associated with infection with SARS-CoV-2 omicron variant versus delta variant in Denmark: an observational cohort study. Lancet Infect Dis 2022; 22:967–76.

11. Nyberg T, Ferguson NM, Nash SG, et al. Comparative analysis of the risks of hospitalisation and death associated with SARS-CoV-2 omicron (B.1.1.529) and delta (B.1.617.2) variants in England: a cohort study. Lancet 2022; 399:1303–12.

12. Taylor CA, Whitaker M, Anglin O, et al. COVID-19-Associated Hospitalizations Among Adults During SARS-CoV-2 Delta and Omicron Variant Predominance, by Race/Ethnicity and Vaccination Status - COVID-NET, 14 States, July 2021-January 2022. MMWR Morb Mortal Wkly Rep 2022; 71:466–73.

13. Skarbinski J, Wood MS, Chervo TC, et al. Risk of severe clinical outcomes among persons with SARS-CoV-2 infection with differing levels of vaccination during widespread Omicron (B.1.1.529) and Delta (B.1.617.2) variant circulation in Northern California: A retrospective cohort study. Lancet Reg Health Am 2022; 12:100297.

14. Harrigan SP, Wilton J, Chong M, et al. Clinical severity of Omicron SARS-CoV-2 variant relative to Delta in British Columbia, Canada: A retrospective analysis of whole genome sequenced cases. Clin Infect Dis 2022; Aug 30;ciac705.

15. Lauring AS, Tenforde MW, Chappell JD, et al. Clinical severity of, and effectiveness of mRNA vaccines against, covid-19 from omicron, delta, and alpha SARS-CoV-2 variants in the United States: prospective observational study. BMJ 2022; 376:e069761.

16. Robinson ML, Morris CP, Betz JF, et al. Impact of SARS-CoV-2 variants on inpatient clinical outcome. Clin Infect Dis 2023 [in press].

17. Beser J, Galanis I, Enkirch T, et al. Seroprevalence of SARS-CoV-2 in Sweden, April 26 to May 9, 2021. Sci Rep 2022; 12:10816.

