## Supplementary material for "Disease severity of unvaccinated SARS-CoV-2 positive adults less than 65 years old without comorbidity, in the Omicron period and pre-Omicron periods": Suppl

### Supplementary materials

#### Supplementary figure legends

**Supplementary Fig. S1.** A) Distribution of SARS-CoV-2 variants over time among whole genome-sequenced cases in Sweden. B) Overall number of study cases per week. C) Study cases hospitalized for COVID-19 per week. Orange represents the Pre-VOC period, green the Alpha period, red the Delta period, blue the Omicron period, and grey mix periods.

**Supplementary Fig S2.** A) Number of weekly SARS-CoV-2-positive individuals aged 18-64 years without comorbidity or care dependency in Sweden according to previous vaccination and previous SARS-CoV-2 positivity. B) Proportion of cases without previous anti-SARS-CoV-2 vaccination and without previous SARS-CoV-2 positivity among SARS-CoV-2-positive individuals aged 18-64 years without comorbidity or care dependency.

**Supplementary Fig. S3.** Odds ratios with 95% confidence intervals for study outcomes among patients hospitalized for COVID-19 after adjusting for age, sex, and socio-economic status with the Delta period as reference.

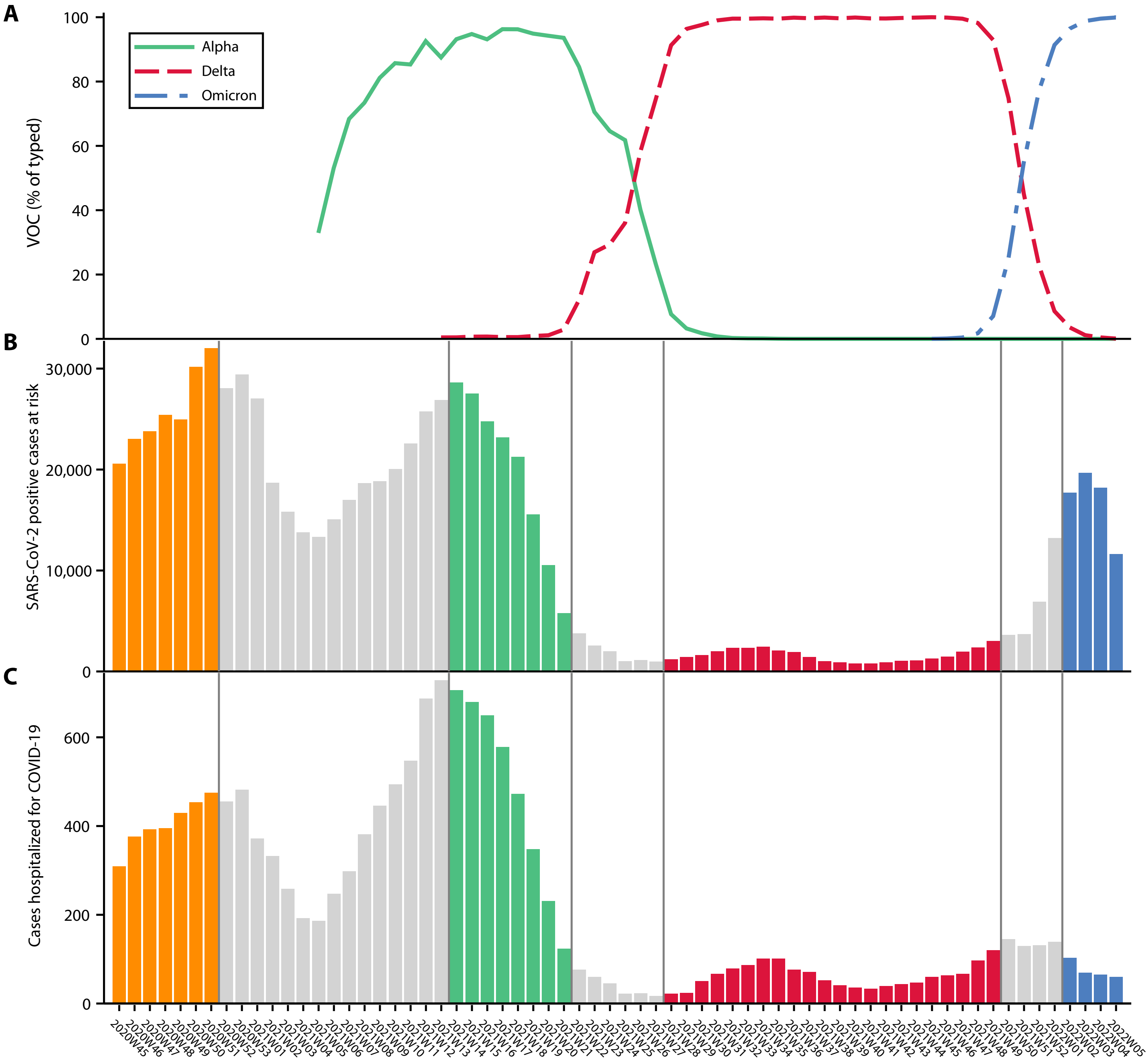
A
B
C
Alpha
Delta
Omicron
VOC (% of typed)
SARS-CoV-2 positive cases at risk
Cases hospitalized for COVID-19
0
20
40
60
80
100
10,000
20,000
30,000
200
400
600
2020W45
2020W46
2020W47
2020W48
2020W49
2020W50
2020W51
2020W52
2020W53
2021W01
2021W02
2021W03
2021W04
2021W05
2021W06
2021W07
2021W08
2021W09
2021W10
2021W11
2021W12
2021W13
2021W14
2021W15
2021W16
2021W17
2021W18
2021W19
2021W20
2021W21
2021W22
2021W23
2021W24
2021W25
2021W26
2021W27
2021W28
2021W29
2021W30
2021W31
2021W32
2021W33
2021W34
2021W35
2021W36
2021W37
2021W38
2021W39
2021W40
2021W41
2021W42
2021W43
2021W44
2021W45
2021W46
2021W47
2021W48
2021W49
2021W50
2021W51
2021W52
2022W01
2022W02
2022W03
2022W04
2022W05

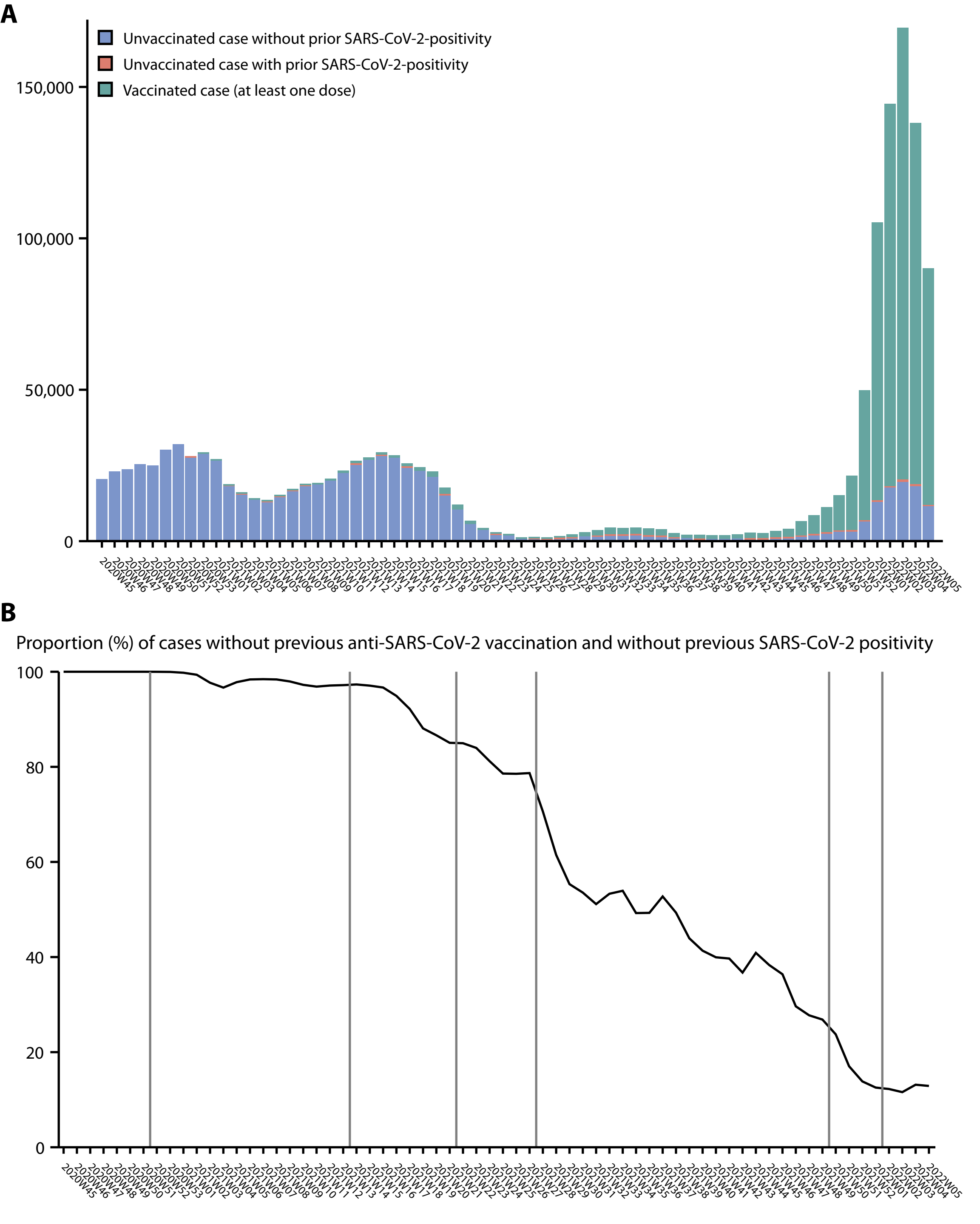

A
Unvaccinated case without prior SARS-CoV-2-positivity
Unvaccinated case with prior SARS-CoV-2-positivity
Vaccinated case (at least one dose)
150,000
100,000
50,000
0
B
Proportion (%) of cases without previous anti-SARS-CoV-2 vaccination and without previous SARS-CoV-2 positivity
100
80
60
40
20
0

**Hospitalized for COVID-19**

Odds Ratio with Delta-period as reference

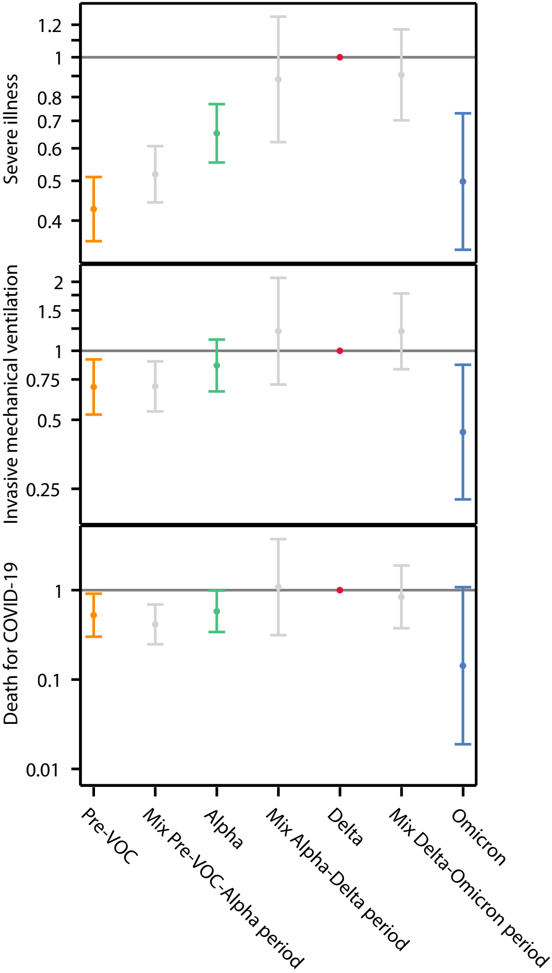
